# Discordance between gut-derived appetite hormones and energy intake in humans

**DOI:** 10.1101/2023.05.10.23289718

**Authors:** Aaron Hengist, Christina M. Sciarrillo, Juen Guo, Mary Walter, Kevin D. Hall

**Affiliations:** National Institute of Diabetes and Digestive and Kidney Diseases, NIH, Bethesda MD, 20892, USA

**Keywords:** Appetite, Energy Intake, Gut Hormones, Postprandial, Low Carbohydrate, Low Fat, Diet

## Abstract

Gut-derived hormones affect appetite and are thought to play an important role in body weight regulation. Dietary macronutrient composition can influence gut-derived appetite hormone concentrations, thereby providing theoretical basis for why some diets might facilitate weight loss better than others. We investigated postprandial gut-derived appetite hormones in 20 inpatient adults after 2 weeks of eating either a low carbohydrate (LC) or a low fat (LF) diet followed by the alternate diet in random order. A LC meal resulted in significantly greater postprandial GLP-1, GIP, and PYY but lower ghrelin compared to an isocaloric LF meal (all *p*≤0.02). However, differences in gut-derived appetite hormones were incommensurate with subsequent ad libitum energy intake over the rest of the day, which was 551±103 kcal (*p*<0.0001) greater with the LC as compared to the LF diet. The effects of gut-derived appetite hormones on ad libitum energy intake can be dominated by other diet-related factors, at least in the short-term.

**Graphical Abstract:** 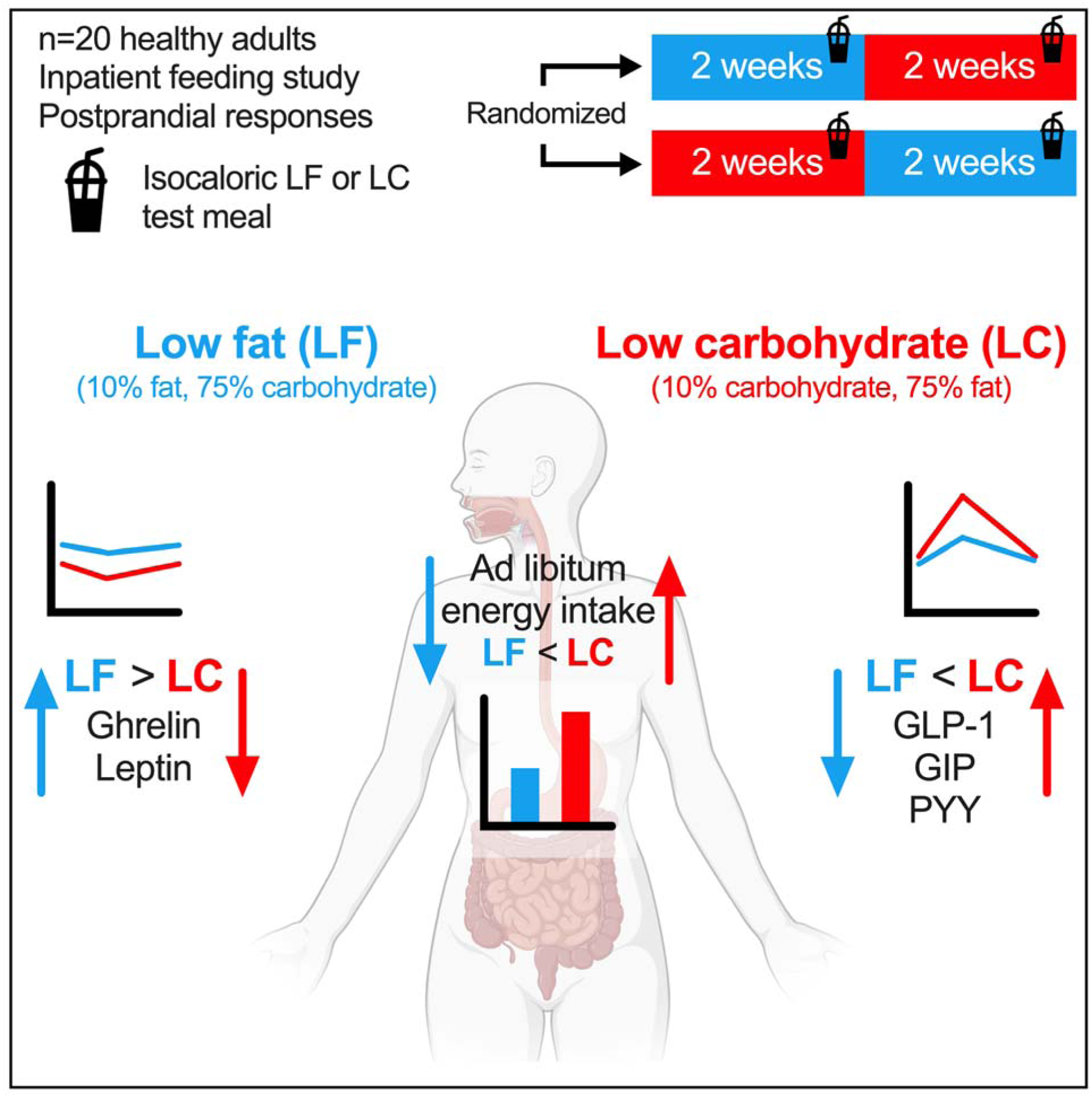

## INTRODUCTION

Gut-derived hormones affect appetite. Ghrelin increases hunger and decreases after food intake, whereas satiation and satiety are induced by peptide YY (PYY), glucagon-like peptide-1 (GLP-1), and perhaps glucose-dependent insulinotropic polypeptide (GIP) which are all increased after food intake.^1-3^ These gut-derived appetite hormones have been theorized to play a role in the weight-loss that results from bariatric surgery^4,5^ and agonists of GLP-1 and GIP receptors have become successful pharmacological treatments for obesity.^6-8^ Circulating concentrations of gut-derived appetite hormones can be influenced by dietary macronutrient composition,^9-13^ which provides a theoretical basis for why some diets may help facilitate weight loss better than others. We recently studied 20 inpatient adults who were exposed to two diets varying widely in the proportion of fat to carbohydrate for periods of 2 weeks each.^14^ In a randomized crossover design, after 2 weeks of eating a low carbohydrate (LC) diet (75% fat, 10% carbohydrate), the postprandial gut hormone responses to a representative LC liquid test meal were compared with an isocaloric low fat (LF) liquid test meal consumed after 2 weeks of eating a LF diet (10% fat, 75% carbohydrate). Subsequent ad libitum energy intake at lunch, dinner, and snacks for the rest of the day were measured to investigate whether postprandial responses were commensurate with subsequent intake during these dietary patterns.

## RESULTS AND DISCUSSION

### Gut hormone responses and subsequent energy intake

At the end of each ad libitum feeding period, the LC diet resulted in greater fasting concentrations of GLP-1 and GIP, but similar concentrations of PYY and leptin, and lower concentrations of total ghrelin and active ghrelin, when compared to the LF diet (**Table 1**). **Figure 1** demonstrates that the LC breakfast test meal delivered at the end of the ad libitum LC period resulted in greater mean postprandial plasma concentrations of active GLP-1 (LC meal: 6.44±0.78 pg·mL^-1^, LF meal: 2.46±0.26 pg·mL^-1^; *p*<0.0001), total GIP (LC meal: 578±60 pg·mL^-1^, LF meal: 319±37 pg·mL^-1^; *p*=0.0002), and PYY (LC meal: 65.6±5.6 pg·mL^-1^, LF meal: 50.7±3.8 pg·mL^-1^; *p*=0.02) whereas total ghrelin (LC meal: 184±25 pg·mL^-1^, LF meal: 261±47 pg·mL^-1^; *p*=0.0009), active ghrelin (LC meal: 91±9 pg·mL^-1^, LF meal: 232±28 pg·mL^-1^; *p*<0.0001), and leptin (LC meal: 26.9±6.5 ng·mL^-1^, LF meal: 35.2±7.5 ng·mL^-1^; *p*=0.01) were lower as compared to an isocaloric LF breakfast test meal delivered at the end of the ad libitum LF period.

**Figure 1.**
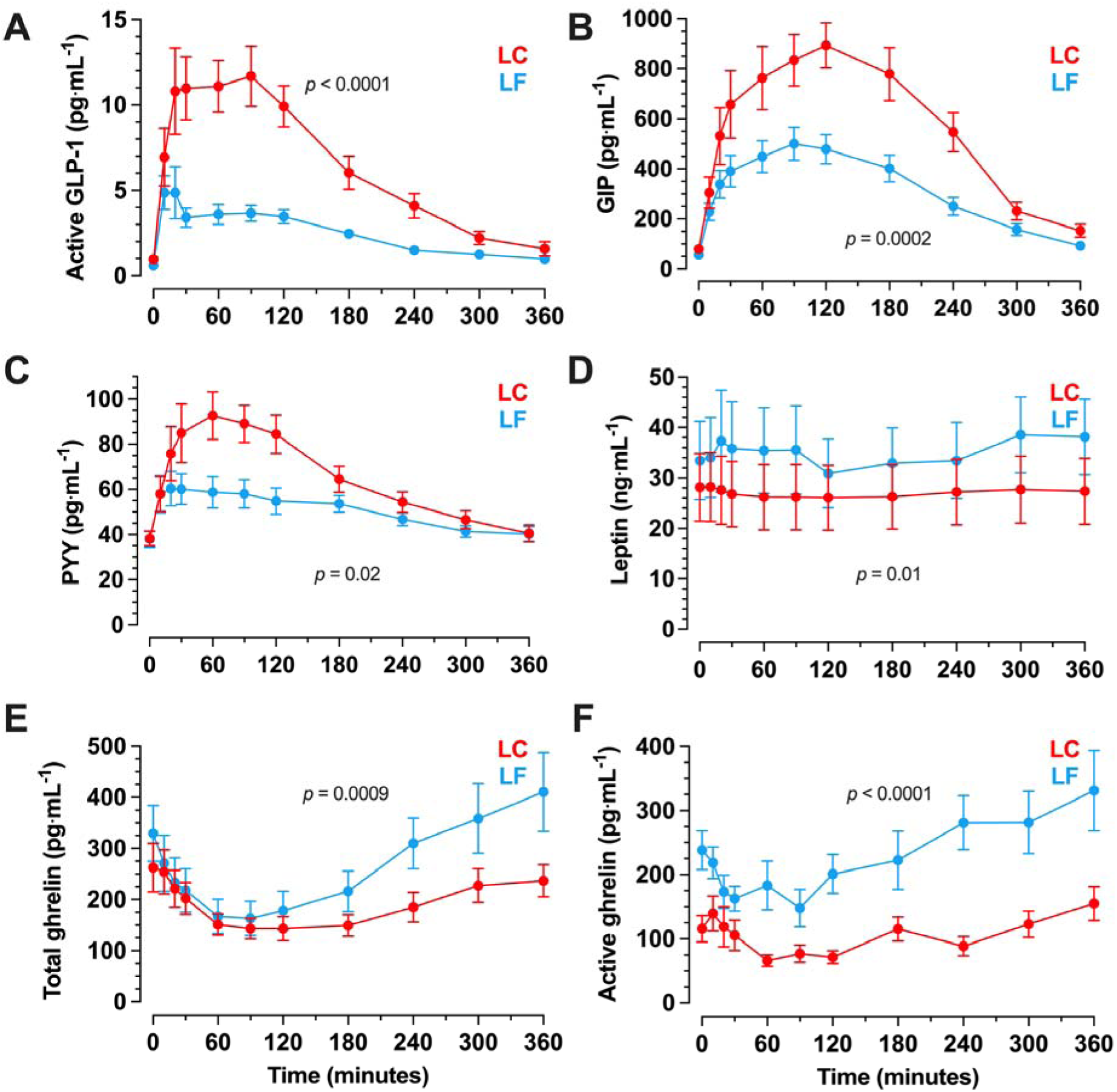
Postprandial responses to isocaloric low carbohydrate (LC) or low fat (LF) meals following habituation to each diet in a randomized crossover design. Mean (range) of energy in the test meals was 777 (532 to 1043) kcal. Data are mean ± SEM. n=20. *p*-values from paired t-test of mean postprandial plasma concentrations. (A) active glucagon-like peptide-1 (GLP-1) (B) total glucose-dependent insulinotropic polypeptide (GIP) (C) peptide YY (PYY) (D) leptin (E) total ghrelin (F) active ghrelin

**Table 1.** Fasting concentrations of gut-derived appetite hormones and leptin in the second week of low carbohydrate (LC) or low fat (LF) diet. Data are mean ± SEM, n=20.

|  | LC | LF | *p* value<br>LC vs LF |
| --- | --- | --- | --- |
| Active GLP-1<br>(pg·mL<sup>-1</sup>) | 0.96 ± 0.14 | 0.61 ± 0.06 | 0.01 |
| Total GIP<br>(pg·mL<sup>-1</sup>) | 80 ± 13 | 56 ± 8 | 0.03 |
| PYY<br>(pg·mL<sup>-1</sup>) | 38.2 ± 3.2 | 38.0 ± 3.7 | 0.94 |
| Total ghrelin<br>(pg·mL<sup>-1</sup>) | 263 ± 48 | 329 ± 55 | 0.004 |
| Active ghrelin<br>(pg·mL<sup>-1</sup>) | 116 ± 21 | 238 ± 30 | 0.0002 |
| Leptin<br>(ng·mL<sup>-1</sup>) | 28.2 ± 6.7 | 33.5 ± 7.8 | 0.39 |

After the breakfast test meal, ad libitum energy intake was greater during the LC diet at lunch (244±85 kcal; *p*=0.001) and dinner (193±86 kcal; *p*=0.04), but not snacks (114±63 kcal; *p*=0.12), such that the total ad libitum energy intake over the rest of the day was significantly greater (551±103 kcal; *p*<0.0001) as compared to when the same participants consumed the LF diet (**Figure 2A**). Within each diet pattern, there were no significant correlations between subsequent ad libitum energy intake and the mean postprandial active GLP-1 (LC diet: r=-0.1; *p*=0.68, LF diet: r=-0.12; *p*=0.60), total GIP (LC diet: r=-0.08; *p*=0.73, LF diet: r=0.23; *p*=0.34), PYY (LC diet: r=-0.23; *p*=0.32, LF diet: r=-0.007; *p*=0.98), total ghrelin (LC diet: r=0.31; *p*=0.19, LF diet: r=0.32; *p*=0.16), active ghrelin (LC diet: r=0.20; *p*=0.41, LF diet: r=0.07; *p*=0.76), or leptin (LC diet: r=-0.07; *p*=0.77, LF diet: r=-0.26; *p*=0.27).

**Figure 2.**
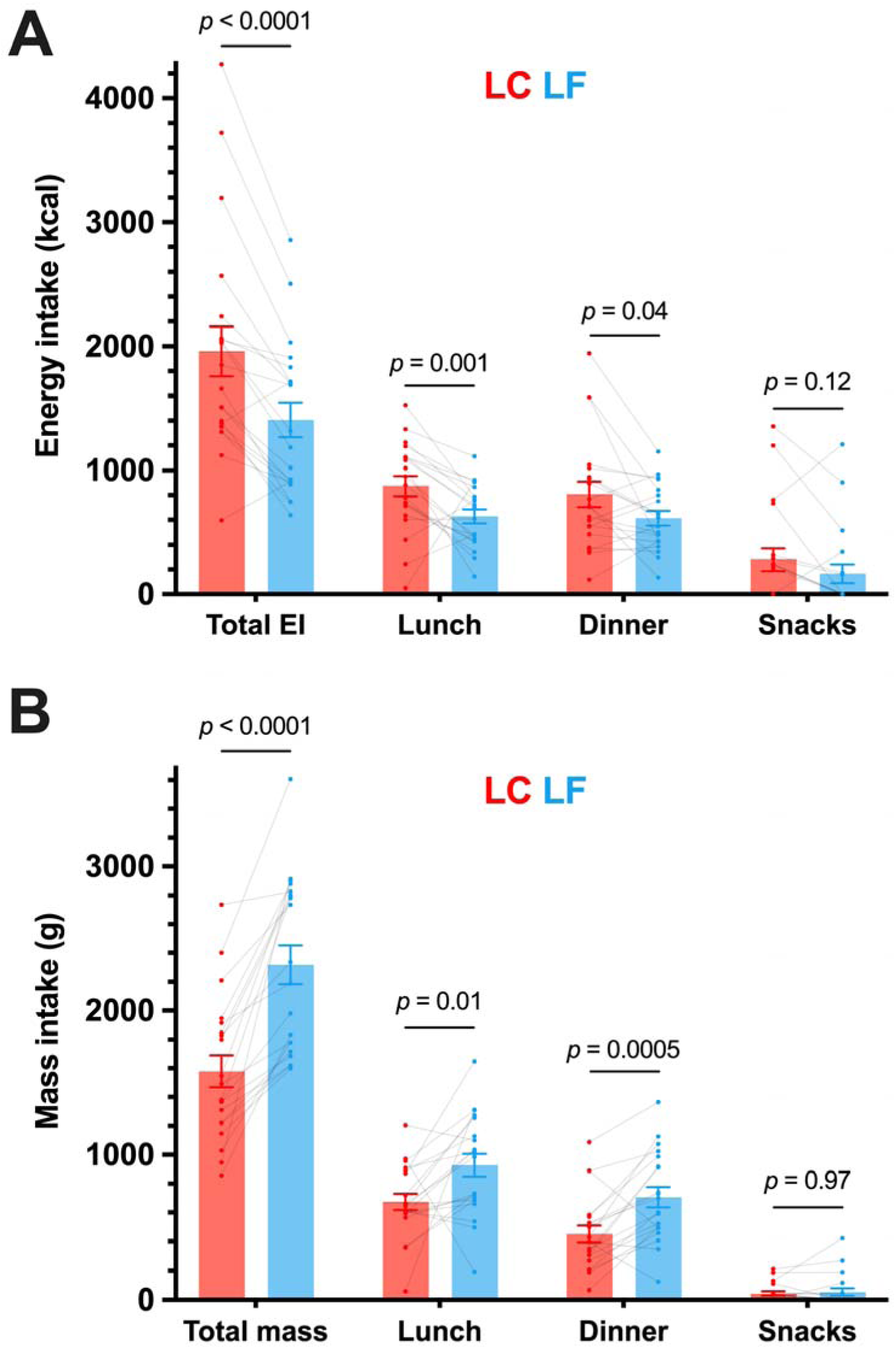
Total intake, and intake from lunch, dinner, and snacks throughout the day after isocaloric low carbohydrate (LC) or low fat (LF) meals following habituation to each diet in a randomized crossover design. Mean (range) of energy in the test meals was 777 (532 to 1043) kcal. Data are mean ± SEM and individual responses. n=20. *p*-values from paired t-test or Wilcoxon test (A) Energy intake (EI) (kcal) (B) Mass intake (g)

The present cohort had a wide range of body mass indices, therefore diet by BMI interactions were explored to investigate if any of the differences in gut hormone responses were driven by BMI (**Figure S1**). The only significant diet by BMI interaction was observed for active ghrelin, which was greater in LF than LC in individuals with a BMI below 25 kg.m^-2^ (*p*<0.0001), but was not siginficiantly different in individuals with overweight (*p*=0.48) or obesity (*p*=0.25).

The observed differences in gut-derived appetite hormones during the LC test meal would be expected to result in reduced appetite and lower ad libitum energy intake as compared to the LF diet. However, the opposite result was observed, with the LC diet resulting in an additional ∼500 kcal consumed over the remainder of the day following the test meal, as compared to the LF diet. This difference was similar to the overall ∼700 kcal·d^-1^ difference between the diets averaged over two weeks.^14^

Concentrations of the adipokine leptin were commensurate with the direction of ad libitum energy intake differences for the LC and LF diets. The lower leptin following the LC diet (*vs* LF) is in agreement with previous evidence comparing a ketogenic low carbohydrate diet with an isocaloric diet moderate in both carbohydrate and fat.^15^ The decrease in leptin following LC is likely explained by decreased insulin and glucose concentrations, which were lower in LC compared to LF.^14^ Previous studies have shown that small increases in insulin induced by glucose infusion of 2.5 mg·kg^-1^·min were sufficient to offset the decrease in leptin observed with 16 hours of fasting^16^ and 24 hours of ketogenic carbohydrate restriction reduced leptin concentrations independent from changes in ad libitum energy intake and preceding changes in adiposity.^17^ Therefore, the evidence suggests that changes in carbohydrate availability, rather than energy intake or energy balance, are key for altering leptin concentrations. Decreased leptin would theoretically increase appetite, as has been associated following weight loss.^18^ Whilst it is possible that leptin, as a longer-term appetite signal, overrides the transient signals from gut-derived hormones, leptin concentrations did not correlate with ad libitum energy intake in the present study, so it is likely that other diet-related factors are more important in this context.

### Influence of macronutrient composition on gut hormone response

Although the LC and LF diets differed by more than just their macronutrient composition, it is likely that the differences in gut hormone responses were mainly due to macronutrient differences as previously reviewed.^19,20^ Early evidence in humans suggested that small increases in GLP-1 were observed after isocaloric carbohydrate (glucose), fat (double cream), or protein (lean turkey) ingestion, whereas GIP only responded to carbohydrate and fat.^21^ However, regardless of nutrient, the food matrix also plays a large role in determining postprandial responses as demonstrated by isocaloric ingestion of glucose eliciting greater GLP-1 and GIP responses than whole food sources of carbohydrate, including brown rice or pearl barley.^21^ With regards to carbohydrate manipulation, the increase in PYY observed following LC in the present study resembles the results of a similar randomized crossover study in participants with obesity who consumed isocaloric low-carbohydrate or low-fat diets for one week before ingesting a representative breakfast meal.^22^ Similarly, high-fat drinks (38% carbohydrate, 50% fat) increase postprandial GLP-1 and PYY responses, without differences in postprandial ghrelin responses, compared to isocaloric (590 kcal) low-fat, high-carbohydrate drinks (84% carbohydrate, 3% fat), but these differences did not translate into differences in ad libitum energy intake in a subsequent lunch meal.^10^ Instead, ad libitum intake was associated with ghrelin responses, which contrasts with our results because total and active ghrelin were reduced with the LC diet in comparison to the LF diet and did not correlate with energy intake.

Macronutrient manipulation, with food volume and energy density controlled, has been shown to alter postprandial GLP-1, GIP, PYY, active ghrelin, and total ghrelin responses, but did not alter subjective hunger or subsequent energy intake.^9,12,13^ Over the course of our study, the LF diet resulted in ∼700 kcal·d^-1^ less ad libitum energy intake as compared to the LC diet without significant differences in self-reported appetite.^14^ Because postprandial responses of gut-derived appetite hormones depend on the amount of food consumed,^23,24^ the expected diet differences in postprandial ghrelin, GLP-1, GIP, and PYY during the ad libitum period would likely be even greater than we observed following the isocaloric meal tests and further emphasizes that the expected effects of these appetite hormone differences were dominated by other diet differences.

### Diet-related factors affecting energy intake beyond gut hormones

Recent analysis of the meal characteristics that affect energy intake from our inpatient feeding studies suggests that energy density, eating rate, and proportion of hyper-palatable foods are positively associated with meal energy intake.^25^ Greater dietary energy density has consistently been shown to increase energy intake in short-term interventions.^26^ The LC diet of the present study had about double the energy density of the LF diet and mediated around 25% of the diet effect on meal energy intake.^25^ A quicker eating rate increases energy intake of presented meals without altering subsequent hunger.^27^ Eating rate could be related to sensory and physical properties of foods, like food texture;^28^ for example, softer, less solid, less viscous foods are associated with increased eating rate.^29^ Eating rate in grams per minute was quicker in the LF meals at lunch (29±9 g·min^-1^, *p*<0.0001) and dinner (14±9 g·min^-1^, *p*=0.009) on the test meal day, compared to LC, so this factor is unlikely to explain our observations of increased energy intake on the LC test meal day. The volume and mass of food ingested is closely related to energy density, which may alter gastric distension and contribute to changes in gut hormone responses to meals.^30^ The mass of food eaten ad libitum was significantly greater following the LF diet at lunch and dinner, but not snacks (**Figure 2B**), compared to LC. This total difference across the day was consistent with the overall ∼667 g·d^-1^ difference between the diets averaged over two weeks.^14^ Within each diet pattern, there were no significant correlations between postprandial gut hormone responses following the liquid test meals and subsequent mass of food eaten at lunch (first subsequent meal) or throughout the day of the test meal (**Table S1**), apart from a moderate negative correlation between food mass intake at lunch in the LF diet and leptin. Hyper-palatable foods have recently been defined using quantitative thresholds of nutrient combinations that may drive excess intake; 1) fat and sugar (>20% energy, >20% energy), 2) fat and sodium (>20% of energy, >0.3% by weight), and 3) carbohydrates and sodium (>40% energy, >0.3% by weight).^31^ Across the entire 2 weeks, meals in the LC diet had a greater proportion of hyper-palatable foods than the LF diet, which may have mediated around 14% of the diet effect on meal energy intake.^25^ The availability of hyperpalatable foods in the US food system, by this definition, has increased from around 49% to around 69% in 30 years.^32^ Emerging cross-sectional evidence suggests that hyper-palatable foods may be more rewarding.^33^ More work is required to determine the utility of quantitative definitions of hyper-palatability and their influence on food intake. Whilst the alternative diet-related factors discussed may often be inter-related in real world settings, it is important for future work to isolate these diet-related factors and test their contribution to ad libitum energy intake in different dietary contexts (e.g. macronutrient manipulation or processing).

### Comparisons between diet and pharmacological or surgical interventions

Discordance between gut hormone responses and energy intake in the present study may appear to contradict the recent success of pharmacological gut hormone mimetics, including GLP-1 receptor agonists, but quantitative considerations of dose and exposure reconcile our findings. Specifically, the estimated active GLP-1 steady state average exposure concentration, C_avg_, for the present study had mean (95% CI) values of 0.034 (0.029, 0.043) nmol·L^-1^ for LF and 0.086 (0.071, 0.113) nmol·L^-1^ for LC, which are orders of magnitude lower than the C_avg_ of both oral and subcutaneous semaglutide ranging from ∼3 nmol·L^-1^ up to ∼30 nmol·L^-1^ with oral and subcutaneous semaglutide, respectively (**Figure 3A**).^34^ Such high C_avg_ with pharmacological treatment is due to the long half-life of semaglutide which has a similar binding affinity to the GLP-1 receptor as native GLP-1,^35^ whereas the half-life of endogenous GLP-1 and GIP is minutes in humans.^36^ Unlike pharmacological intervention, diet-induced changes in gut hormone concentrations reflect conjoint changes of multiple hormones in a complex signalling system, so the quantitative exposure of GLP-1 from diet and pharmacological interventions cannot be compared directly, but this comparison highlights that the magnitude of change in GLP-1 from diet is not comparable to that of pharmacological interventions even with the concurrent changes of other hormones.

**Figure 3.**
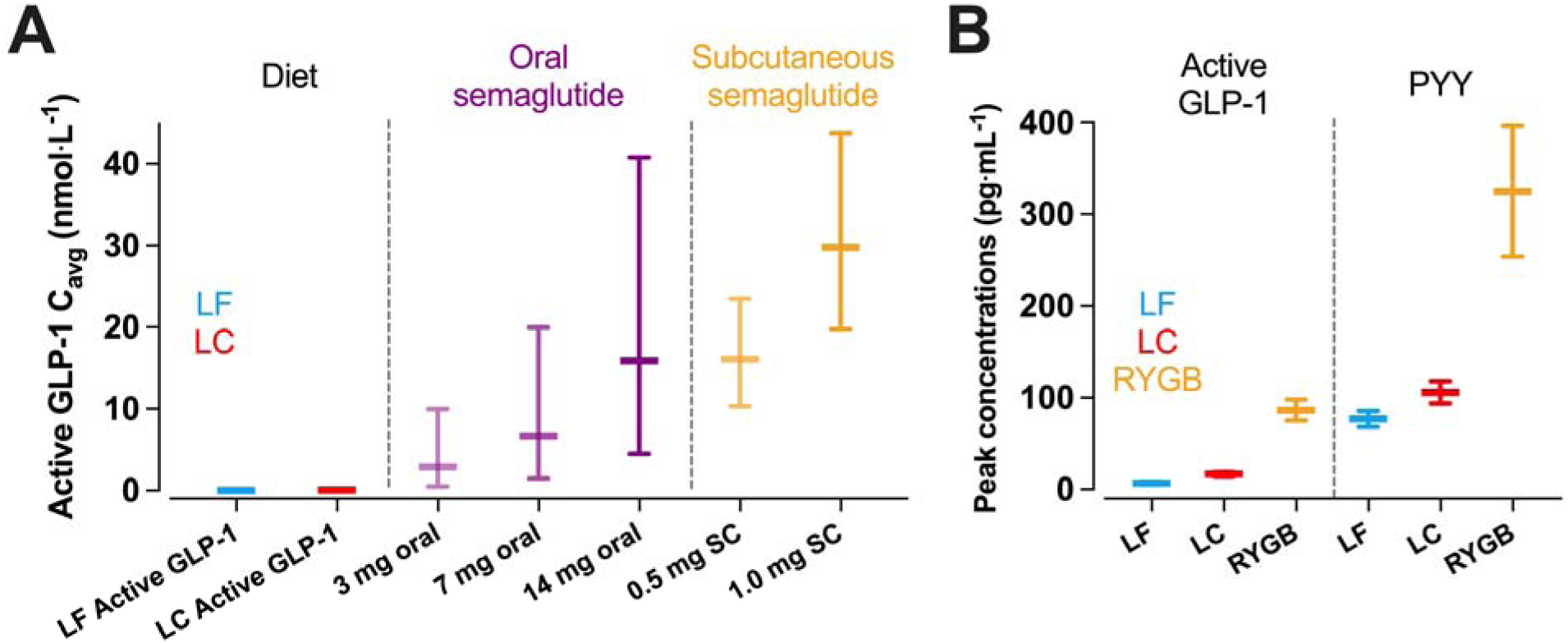
Comparisons between dietary macronutrient induced changes in gut hormone responses and pharmacological or bariatric surgery induced changes. (A) estimated mean active GLP-1 steady state average exposure concentrations, C_avg_, achieved by low carbohydrate (LC) or low fat (LF) diet were orders of magnitude lower than the both oral and subcutaneous semaglutide using values median (90% exposure ranges) from Overgaard et al.^34^ (B) peak active GLP-1 and PYY concentrations following a LC or LF test meal were orders of magnitude lower than peak concentrations observed during a mixed-meal test following Roux-en-Y Gastric Bypass surgery (RYGB) using data from Tan et al.^3^ Data are mean ± SEM.

Pharmacological engagement of the GLP-1 receptor (e.g. exogenous GLP-1) may act differently from nutrient-stimulated gut hormones (e.g. endogenous GLP-1). For example, endogenous GLP-1 may affect the hypothalamus through neuronal signaling from the gut to brain via solitary tract neurons, whereas exogenous GLP-1 receptor agonists may directly engage hypothalamic and brainstem GLP-1 receptors.^6^ Further complicating this, GLP-1 can be produced centrally in brain regions which may directly alter appetite independent from diet-induced gut hormone secretion.^37^ With this context, changes in endogenous gut hormone concentrations induced by diet may be not be potent enough to affect energy intake. Indeed, mouse models that knockout the GLP-1 receptor,^38^ or delete intestinal GLP-1 production throught GNG gene knockout,^39^ do not result in a body weight or food intake phenotype, suggesting that endogenous GLP-1 has a limited effect on appetite in the normal physiological range.

Some forms of bariatric surgery result in substantial increases in postprandial GLP-1 and PYY,^40-42^ likely due to altered gastric emptying and intestinal nutrient delivery. The magnitude of post-surgical changes in postprandial gut hormone responses might be mechanistically linked to reduced appetite and energy intake. Infusion of GLP-1, PYY, and oxyntomodulin in healthy participants that mimics the concentrations observed following Roux-en-Y gastric bypass reduced energy intake at lunch and dinner by ∼400 kcal.^3^ The active GLP-1 and PYY concentrations achieved were around 26 pmol·L^-1^ (85 pg·mL^-1^) and 80 pmol·L^-1^ (320 pg·mL^-1^) respectively. For active GLP-1, these concentrations are around 13-fold and 35-fold greater than mean postprandial concentrations following the LC and LF meals in the present study, and for PYY they are around 5-fold and 6-fold greater (**Figure 3B**). Infusing GLP-1 to achieve concentrations comparable with the LC condition of the present study (∼15 pg·mL^-1^) delays gastric emptying without suppressing subjective appetite and ad libitum intake.^43^ Supraphysiological concentrations of GLP-1 achieved by infusion (∼25 to 30 pg·mL^-1^) suppress subjective appetite, but effects on subsequent ad libitum energy intake are modest compared with higher concentrations (∼100 to 240 kcal).^44,45^ Together, infusion studies indicate a dose-response relationship between active GLP-1 and suppression of appetite,^46^ and suggest that substantially greater increases in gut hormone concentrations are required to have meaningful effects on appetite and energy intake, likely greater than is achievable by diet interventions alone.

### Limitations and considerations

A limitation of the current study is that participants had no choice regarding the foods available for consumption. They could only choose the quantity of the foods eaten. While the gut-derived appetite hormones were not a dominant factor determining energy intake in this setting, it is possible that such differences in appetite hormones in a real-world setting might alter food choices at subsequent meals and thereby alter energy intake. Another limitation is that we used isocaloric liquid test meals that matched the macronutrient composition of the overall LC and LF diets. These test meals may have not been adequately representative of the effects of meals with solid foods. Furthermore, the results reported in this study were from analyses that were not pre-specified as primary or secondary outcomes of the main study and are hence exploratory in nature. Nevertheless, our study suggests that differences in dietary factors like energy density or proportion of hyper-palatable foods may play a greater role in appetite regulation than endogenous gut-derived appetite hormones, at least in the short term. Future research should aim to identify such diet differences that influence energy intake and evaluate whether their effects and their potential discordance with gut-dervied appetite hormones persist over time.

## Data Availability

Deidentified individual subject data from consenting participants will be posted at https://osf.io/fjykq/

## ACKNOWLEDGEMENTS

This work was supported by the Intramural Research Program of the NIH, National Institute of Diabetes & Digestive & Kidney Diseases under award number 1ZIADK013037. We thank the nursing and nutrition staff at the NIH Metabolic Clinical Research Unit for their invaluable assistance with this study. We thank the study participants for their invaluable contribution.

## AUTHOR CONTRIBUTIONS

KDH designed the study, MW performed the biochemical analyses, JG and KDH analyzed the data, AH, CMS, JG, and KDH interpreted the data, drafted the manuscript, and approved the final version.

## STAR Methods

### KEY REOURCES TABLE

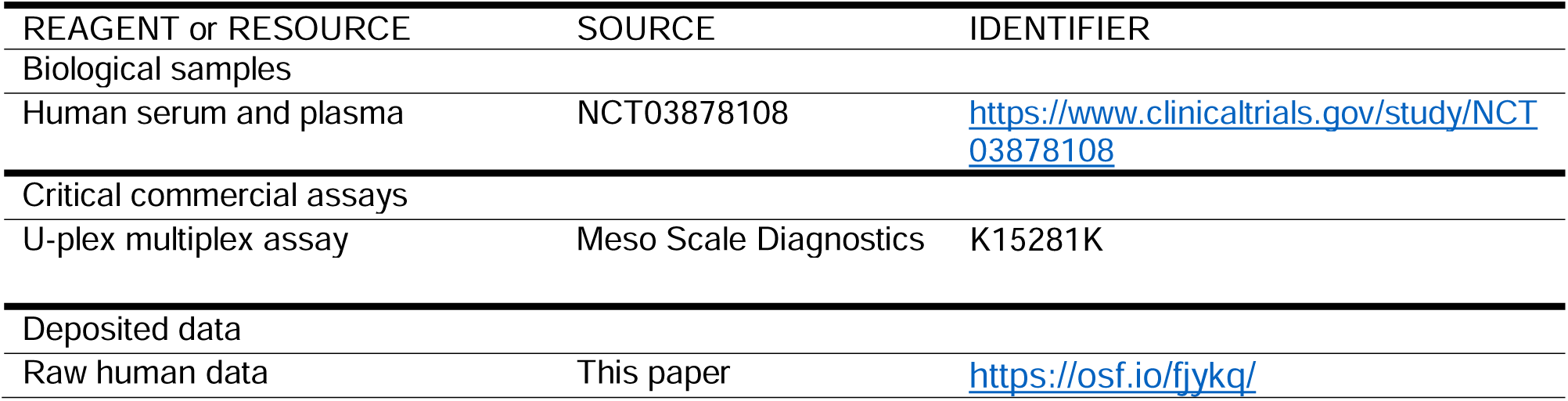

### RESOURCE AVAILABILITY

#### Lead contact

Further information and requests for resources and reagents should be directed to and will be fulfilled by the Lead Contact Dr Kevin Hall.

#### Materials availability

This study did not generate new unique reagents.

#### Data and code availability

De-identified data have been deposited at https://osf.io/fjykq/ and are publicly available as of the date of publication. This paper does not report original code.

### EXPERIMENTAL MODEL AND STUDY PARTICIPANT DETAILS

These data are exploratory endpoints from a registered clinical trial conducted at the Metabolic Clinical Research Unit at the NIH Clinical Center and approved by the institutional board of the National Institute of Diabetes & Digestive & Kidney Diseases (NCT03878108). Detailed methods have been published previously.^14^ Participants completed a single four week stay and completed 2 diets in a randomized order. One diet was an animal-based, ketogenic, low carbohydrate (LC) diet with ∼10% of energy from carbohydrates, ∼75% from fat and the other diet was a plant-based, low fat (LF) diet with ∼10% of energy from fat, ∼75% from carbohydrate. Inclusion criteria were male and female adults age 18–50 years; weight stable (within 5% in past 6 months); body mass index ≥20Dkg·m^-2^; body weight ≥53 kg; able to complete daily bouts of stationary cycling at a moderate rate and intensity with a heart rate (HR) equal to or greater than 0.3D×D(220D−DageD−DHRrest)D+DHRrest but not exceeding 0.4D×D(220D−DageD−DHRrest)D+DHRrest and no signs of arrhythmia. Exclusion criteria were evidence of metabolic or cardiovascular disease or disease that may influence metabolism (for example cancer, diabetes, thyroid disease); taking any prescription medication or other drug that may influence metabolism (for example diet/weight-loss medication, asthma medication, blood pressure medication, psychiatric medications, corticosteroids or other medications at the discretion of the study team); positive pregnancy test or lactation as determined by volunteer report (women only); participating in a regular exercise program (>2DhDweek−1 of vigorous activity); hematocrit <37% for women and <40% for men; habitual caffeine consumption >300DmgDd−1; regular use of alcohol (>2 drinks per day), tobacco (smoking or chewing), amphetamines, cocaine, heroin or marijuana over past 6 months; psychological conditions such as (but not limited to) eating disorders, claustrophobia, clinical depression, bipolar disorders, as determined by investigators after reviewing the results of the DSM-5 Self-Rated Level 1 Cross-Cutting Symptom Measure; past or present history of claustrophobia; implants, devices or foreign objects implanted in the body that interfere with the magnetic resonance procedures; strict dietary concerns (for example vegetarian or kosher diet, food allergies) as determined by investigators after reviewing the results of the Food Frequency Questionnaire; volunteers unwilling or unable to give informed consent; and non-English speakers owing to unavailability of required questionnaires in other languages. Participants were 20 adults (male n=11, female n=9; mean±SD, age 30±6 years; body mass 80.8±18.2 kg; body mass index 27.8±5.9 kg·m^-2^; fat mass 26.9±11.2 kg; body fat percentage 32.8±9.8%; resting energy expenditure 1550±287 kcal·d^-1^).

### METHOD DETAILS

At the end of the second inpatient week of consuming either the ad libitum low carbohydrate (LC) or low fat (LF) diets and after an overnight fast, participants consumed liquid meals matching the macronutrient content of the prevailing diet and amounting to 30% of the estimated daily calorie requirements as determined by 1.6 multiplied by the resting energy expenditure measured at screening. Mean (range) of energy in the test meals was 777 (532 to 1043) kcal. The LC test meal was 10% carbohydrate, 75% fat, and 15% protein whereas the LF test meal was 75% carbohydrate, 10% fat, and 15% protein. Blood samples were obtained at 0, 10, 20, 30, 60, 90, 120, 180, 240, 300, and 360 minutes after the meals in tubes containing a protease inhibitor cocktail (including DPPIV inhibitor and aprotinin) to measure GLP-1, GIP, PYY, total ghrelin, active ghrelin, and leptin using multiplex immunoassays (Meso Scale Diagnostics).

After the breakfast mixed meal tests, ad libitum food intake was measured over the rest of the day including lunch, dinner, and snacks by weighing the remaining food and beverages to calculate the amount of each food consumed and energy intake was calculated using ProNutra software (v.3.4, Viocare) with nutrient values derived from the USDA National Nutrient Database for Standard Reference, Release 26 and the USDA Food and Nutrient Database for Dietary Studies, 4.0. Statistical analyses were performed using SAS (v.9.4; SAS Institute) and Prism (v.9.5.0; GraphPad). Mean plasma concentrations were calculated by dividing total area under the curve (tAUC) by 360 minutes. Active GLP-1 C_avg_ was estimated by multiplying the 6-hour postprandial tAUC by 3 (18 h) and multiplying the postabsorptive (fasting) concentration by 360 minutes (6 h), to get 24-hour exposure, and dividing by 24. The conversion factor used for GLP-1 was 1 pmol·L^-1^ = 3.297 pg·mL^-1^. Data were checked for normality using Shapiro-Wilk test, differences between conditions were assessed using paired t-tests for normally distributed and Wilcoxon tests for non-normally distributed data. Simple linear correlation was used to explore associations between gut hormone responses and ad libitum energy intake. Diet by BMI interactions were checked using 2-way ANOVA, with post-hoc Bonferroni tests used to identify differences. Significance was accepted as *p*≤0.05.

## SUPPLEMENTAL INFORMATION

**Figure S1.**
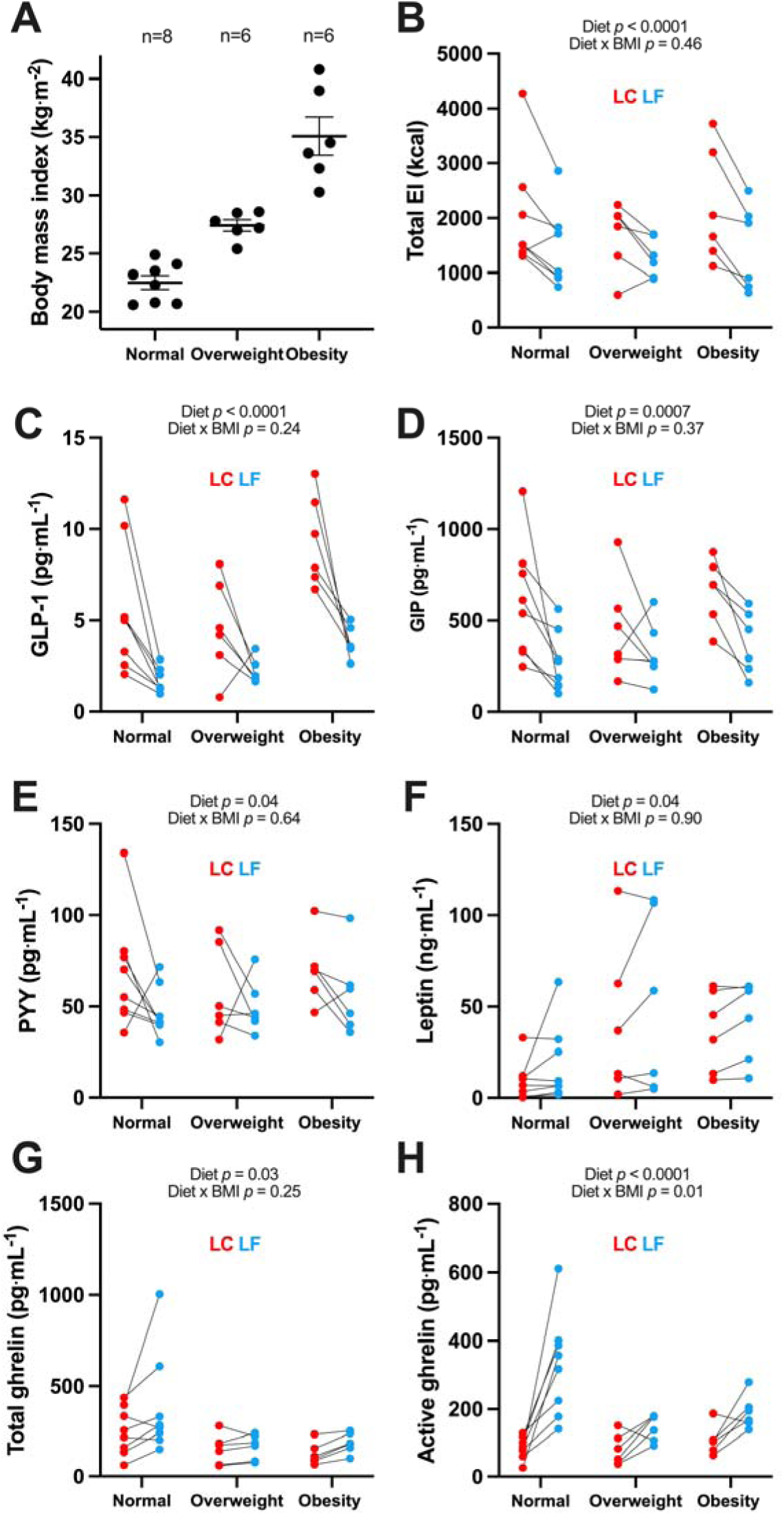
Gut hormone responses sub-divided into body mass index categories. Data are mean ± SEM and individual responses. n=20. *p*-values from two-way ANOVA of diet by BMI category. (A) Participants split by BMI category (B) Total energy intake (C) active glucagon-like peptide-1 (GLP-1) (D) total glucose-dependent insulinotropic polypeptide (GIP) (E) peptide YY (PYY) (F) leptin (G) total ghrelin (H) active ghrelin

**Table S1.** Correlations between mean postprandial responses to isocaloric low carbohydrate (LC) or low fat (LF) meals following habituation to each diet in a randomized crossover design with the subsequent mass intake in the lunch meal and total intake across the day following the test meal.

| Mean<br>postprandial<br>concentrations | Subsequent **lunch**<br>mass intake during test<br>meal day |  | Subsequent **total** mass<br>intake during test meal<br>day |  |
| --- | --- | --- | --- | --- |
|  | LC | LF | LC | LF |
| Active GLP-1 | -0.11<br>(0.63) | -0.21<br>(0.37) | -0.12<br>(0.62) | -0.14<br>(0.55) |
| Total GIP | -0.20<br>(0.39) | -0.06<br>(0.79) | -0.15<br>(0.54) | 0.22<br>(0.36) |
| PYY | -0.22<br>(0.36) | 0.07<br>(0.76) | -0.33<br>(0.15) | 0.10<br>(0.68) |
| Total ghrelin | -0.30<br>(0.20) | 0.44<br>(0.0504) | -0.06<br>(0.82) | 0.38<br>(0.10) |
| Active ghrelin | 0.004<br>(0.99) | 0.35<br>(0.13) | 0.06<br>(0.81) | 0.20<br>(0.39) |
| Leptin | -0.06<br>(0.81) | **-0.48**<br>**(0.03)** | -0.07<br>(0.76) | -0.40<br>(0.08) |

Data are Pearson's r (p-value). n=20.

## REFERENCES

1. Samms, R.J., Goghlan, M.P., and Sloop, K.W. (2020). How May GIP Enhance the Therapeutic Efficacy of GLP-1? Trends in Endocrinology and Metabolism 31, 410–421. 10.1016/j.tem.2020.02.006.

2. Sato, T., Nakamura, Y., Shiimura, Y., Ohgusu, H., Kangawa, K., and Kojima, M. (2012). Structure, regulation and function of ghrelin. J Biochem 151, 119–128. 10.1093/jb/mvr134.

3. Tan, T., Behary, P., Tharakan, G., Minnion, J., Al-Najim, W., Albrechtsen, N.J.W., Holst, J.J., and Bloom, S.R. (2017). The Effect of a Subcutaneous Infusion of GLP-1, OXM, and PYY on Energy Intake and Expenditure in Obese Volunteers. The Journal of clinical endocrinology and metabolism 102, 2364–2372. 10.1210/jc.2017-00469.

4. Holst, J.J., Madsbad, S., Bojsen-Moller, K.N., Svane, M.S., Jorgensen, N.B., Dirksen, C., and Martinussen, C. (2018). Mechanisms in bariatric surgery: Gut hormones, diabetes resolution, and weight loss. Surg Obes Relat Dis 14, 708–714. 10.1016/j.soard.2018.03.003.

5. Troke, R.C., Tan, T.M., and Bloom, S.R. (2014). The future role of gut hormones in the treatment of obesity. Ther Adv Chronic Dis 5, 4–14. 10.1177/2040622313506730.

6. Drucker, D.J., and Holst, J.J. (2023). The expanding incretin universe: from basic biology to clinical translation. Diabetologia. 10.1007/s00125-023-05906-7.

7. Jastreboff, A.M., Aronne, L.J., Ahmad, N.N., Wharton, S., Connery, L., Alves, B., Kiyosue, A., Zhang, S., Liu, B., Bunck, M.C., et al. (2022). Tirzepatide Once Weekly for the Treatment of Obesity. N Engl J Med 387, 205–216. 10.1056/NEJMoa2206038.

8. Wilding, J.P.H., Batterham, R.L., Calanna, S., Davies, M., Van Gaal, L.F., Lingvay, I., McGowan, B.M., Rosenstock, J., Tran, M.T.D., Wadden, T.A., et al. (2021). Once-Weekly Semaglutide in Adults with Overweight or Obesity. N Engl J Med 384, 989–1002. 10.1056/NEJMoa2032183.

9. Foster-Schubert, K.E., Overduin, J., Prudom, C.E., Liu, J., Callahan, H.S., Gaylinn, B.D., Thorner, M.O., and Cummings, D.E. (2008). Acyl and total ghrelin are suppressed strongly by ingested proteins, weakly by lipids, and biphasically by carbohydrates. The Journal of clinical endocrinology and metabolism 93, 1971–1979. 10.1210/jc.2007-2289.

10. Gibbons, C., Caudwell, P., Finlayson, G., Webb, D.L., Hellstrom, P.M., Naslund, E., and Blundell, J.E. (2013). Comparison of postprandial profiles of ghrelin, active GLP-1, and total PYY to meals varying in fat and carbohydrate and their association with hunger and the phases of satiety. The Journal of clinical endocrinology and metabolism 98, E847–855. 10.1210/jc.2012-3835.

11. Helou, N., Obeid, O., Azar, S.T., and Hwalla, N. (2008). Variation of postprandial PYY 3-36 response following ingestion of differing macronutrient meals in obese females. Ann Nutr Metab 52, 188–195. 10.1159/000138122.

12. Raben, A., Agerholm-Larsen, L., Flint, A., Holst, J.J., and Astrup, A. (2003). Meals with similar energy densities but rich in protein, fat, carbohydrate, or alcohol have different effects on energy expenditure and substrate metabolism but not on appetite and energy intake. Am J Clin Nutr 77, 91–100. DOI 10.1093/ajcn/77.1.91.

13. van der Klaauw, A.A., Keogh, J.M., Henning, E., Trowse, V.M., Dhillo, W.S., Ghatei, M.A., and Farooqi, I.S. (2013). High Protein Intake Stimulates Postprandial GLP1 and PYY Release. Obesity 21, 1602–1607. 10.1002/oby.20154.

14. Hall, K.D., Guo, J., Courville, A.B., Boring, J., Brychta, R., Chen, K.Y., Darcey, V., Forde, C.G., Gharib, A.M., Gallagher, I., et al. (2021). Effect of a plant-based, low-fat diet versus an animal-based, ketogenic diet on ad libitum energy intake. Nat Med 27, 344–353. 10.1038/s41591-020-01209-1.

15. Hall, K.D., Chen, K.Y., Guo, J., Lam, Y.Y., Leibel, R.L., Mayer, L.E., Reitman, M.L., Rosenbaum, M., Smith, S.R., Walsh, B.T., and Ravussin, E. (2016). Energy expenditure and body composition changes after an isocaloric ketogenic diet in overweight and obese men. Am J Clin Nutr 104, 324–333. 10.3945/ajcn.116.133561.

16. Sonnenberg, G.E., Krakower, G.R., Hoffmann, R.G., Maas, D.L., Hennes, M.M., and Kissebah, A.H. (2001). Plasma leptin concentrations during extended fasting and graded glucose infusions: relationships with changes in glucose, insulin, and FFA. The Journal of clinical endocrinology and metabolism 86, 4895–4900. 10.1210/jcem.86.10.7951.

17. Hengist, A., Davies, R.G., Rogers, P.J., Brunstrom, J.M., van Loon, L.J.C., Walhin, J.P., Thompson, D., Koumanov, F., Betts, J.A., and Gonzalez, J.T. (2022). Restricting sugar or carbohydrate intake does not impact physical activity level or energy intake over 24 h despite changes in substrate use: a randomised crossover study in healthy men and women. Eur J Nutr. 10.1007/s00394-022-03048-x.

18. Keim, N.L., Stern, J.S., and Havel, P.J. (1998). Relation between circulating leptin concentrations and appetite during a prolonged, moderate energy deficit in women. Am J Clin Nutr 68, 794–801. 10.1093/ajcn/68.4.794.

19. Bodnaruc, A.M., Prud’homme, D., Blanchet, R., and Giroux, I. (2016). Nutritional modulation of endogenous glucagon-like peptide-1 secretion: a review. Nutr Metab (Lond) 13, 92. 10.1186/s12986-016-0153-3.

20. Watkins, J.D., Koumanov, F., and Gonzalez, J.T. (2021). Protein- and Calcium-Mediated GLP-1 Secretion: A Narrative Review. Adv Nutr 12, 2540–2552. 10.1093/advances/nmab078.

21. Elliott, R.M., Morgan, L.M., Tredger, J.A., Deacon, S., Wright, J., and Marks, V. (1993). Glucagon-like peptide-1 (7-36)amide and glucose-dependent insulinotropic polypeptide secretion in response to nutrient ingestion in man: acute post-prandial and 24-h secretion patterns. J Endocrinol 138, 159–166. 10.1677/joe.0.1380159.

22. Essah, P.A., Levy, J.R., Sistrun, S.N., Kelly, S.M., and Nestler, J.E. (2007). Effect of macronutrient composition on postprandial peptide YY levels. Journal of Clinical Endocrinology & Metabolism 92, 4052–4055. 10.1210/jc.2006-2273.

23. Hengist, A., Edinburgh, R.M., Davies, R.G., Walhin, J.P., Buniam, J., James, L.J., Rogers, P.J., Gonzalez, J.T., and Betts, J.A. (2020). Physiological responses to maximal eating in men. The British journal of nutrition 124, 407–417. 10.1017/S0007114520001270.

24. Lewis, H.B., Ahern, A.L., Solis-Trapala, I., Walker, C.G., Reimann, F., Gribble, F.M., and Jebb, S.A. (2015). Effect of reducing portion size at a compulsory meal on later energy intake, gut hormones, and appetite in overweight adults. Obesity (Silver Spring) 23, 1362–1370. 10.1002/oby.21105.

25. Fazzino, T.L., Courville, A.B., Guo, J., and Hall, K.D. (2023). Ad libitum meal energy intake is positively influenced by energy density, eating rate and hyper-palatable food across four dietary patterns. Nat Food. 10.1038/s43016-022-00688-4.

26. Robinson, E., Khuttan, M., McFarland-Lesser, I., Patel, Z., and Jones, A. (2022). Calorie reformulation: a systematic review and meta-analysis examining the effect of manipulating food energy density on daily energy intake. Int J Behav Nutr Phy 19. ARTN 48 10.1186/s12966-022-01287-z.

27. Robinson, E., Almiron-Roig, E., Rutters, F., de Graaf, C., Forde, C.G., Smith, C.T., Nolan, S.J., and Jebb, S.A. (2014). A systematic review and meta-analysis examining the effect of eating rate on energy intake and hunger. Am J Clin Nutr 100, 123–151. 10.3945/ajcn.113.081745.

28. Forde, C.G., and de Graaf, K. (2022). Influence of Sensory Properties in Moderating Eating Behaviors and Food Intake. Frontiers in Nutrition 9. ARTN 841444 10.3389/fnut.2022.841444.

29. Appleton, K.M., Newbury, A., Almiron-Roig, E., Yeomans, M.R., Brunstrom, J.M., de Graaf, K., Geurts, L., Kildegaard, H., and Vinoy, S. (2021). Sensory and physical characteristics of foods that impact food intake without affecting acceptability: Systematic review and meta-analyses. Obesity Reviews 22. 10.1111/obr.13234.

30. Cummings, D.E., and Overduin, J. (2007). Gastrointestinal regulation of food intake. The Journal of clinical investigation 117, 13–23. 10.1172/JCI30227.

31. Fazzino, T.L., Rohde, K., and Sullivan, D.K. (2019). Hyper-Palatable Foods: Development of a Quantitative Definition and Application to the US Food System Database. Obesity 27, 1761–1768. 10.1002/oby.22639.

32. Demeke, S., Rohde, K., Chollet-Hinton, L., Sutton, C., Kong, K.L., and Fazzino, T.L. (2023). Change in hyper-palatable food availability in the US food system over 30 years: 1988-2018. Public health nutrition 26, 182–189. Pii S1368980022001227 10.1017/S1368980022001227.

33. Fazzino, T.L., Bjorlie, K., Rohde, K., Smith, A., and Yi, R. (2022). Choices Between Money and Hyper-Palatable Food: Choice Impulsivity and Eating Behavior. Health Psychol 41, 538–548. 10.1037/hea0001185.

34. Overgaard, R.V., Hertz, C.L., Ingwersen, S.H., Navarria, A., and Drucker, D.J. (2021). Levels of circulating semaglutide determine reductions in HbA1c and body weight in people with type 2 diabetes. Cell Rep Med 2, 100387. 10.1016/j.xcrm.2021.100387.

35. Overgaard, R.V., Navarria, A., Ingwersen, S.H., Baekdal, T.A., and Kildemoes, R.J. (2021). Clinical Pharmacokinetics of Oral Semaglutide: Analyses of Data from Clinical Pharmacology Trials. Clin Pharmacokinet 60, 1335–1348. 10.1007/s40262-021-01025-x.

36. Meier, J.J., Nauck, M.A., Kranz, D., Holst, J.J., Deacon, C.F., Gaeckler, D., Schmidt, W.E., and Gallwitz, B. (2004). Secretion, degradation, and elimination of glucagon-like peptide 1 and gastric inhibitory polypeptide in patients with chronic renal insufficiency and healthy control subjects. Diabetes 53, 654–662. 10.2337/diabetes.53.3.654.

37. Daniels, D., and Mietlicki-Baase, E.G. (2019). Glucagon-Like Peptide 1 in the Brain: Where Is It Coming From, Where Is It Going? Diabetes 68, 15–17. 10.2337/dbi18-0045.

38. Scrocchi, L.A., Brown, T.J., MaClusky, N., Brubaker, P.L., Auerbach, A.B., Joyner, A.L., and Drucker, D.J. (1996). Glucose intolerance but normal satiety in mice with a null mutation in the glucagon-like peptide 1 receptor gene. Nat Med 2, 1254–1258. 10.1038/nm1196-1254.

39. Song, Y., Koehler, J.A., Baggio, L.L., Powers, A.C., Sandoval, D.A., and Drucker, D.J. (2019). Gut-Proglucagon-Derived Peptides Are Essential for Regulating Glucose Homeostasis in Mice. Cell Metab 30, 976–986 e973. 10.1016/j.cmet.2019.08.009.

40. Lampropoulos, C., Alexandrides, T., Tsochatzis, S., Kehagias, D., and Kehagias, I. (2021). Are the Changes in Gastrointestinal Hormone Secretion Necessary for the Success of Bariatric Surgery? A Critical Review of the Literature. Obes Surg 31, 4575–4584. 10.1007/s11695-021-05568-7.

41. Papamargaritis, D., and le Roux, C.W. (2021). Do Gut Hormones Contribute to Weight Loss and Glycaemic Outcomes after Bariatric Surgery? Nutrients 13. 10.3390/nu13030762.

42. Yousseif, A., Emmanuel, J., Karra, E., Millet, Q., Elkalaawy, M., Jenkinson, A.D., Hashemi, M., Adamo, M., Finer, N., Fiennes, A.G., et al. (2014). Differential effects of laparoscopic sleeve gastrectomy and laparoscopic gastric bypass on appetite, circulating acyl-ghrelin, peptide YY3-36 and active GLP-1 levels in non-diabetic humans. Obes Surg 24, 241–252. 10.1007/s11695-013-1066-0.

43. Flint, A., Raben, A., Ersboll, A.K., Holst, J.J., and Astrup, A. (2001). The effect of physiological levels of glucagon-like peptide-1 on appetite, gastric emptying, energy and substrate metabolism in obesity. International journal of obesity and related metabolic disorders: journal of the International Association for the Study of Obesity 25, 781–792. 10.1038/sj.ijo.0801627.

44. Flint, A., Raben, A., Astrup, A., and Holst, J.J. (1998). Glucagon-like peptide 1 promotes satiety and suppresses energy intake in humans. The Journal of clinical investigation 101, 515–520. 10.1172/JCI990.

45. Long, S.J., Sutton, J.A., Amaee, W.B., Giouvanoudi, A., Spyrou, N.M., Rogers, P.J., and Morgan, L.M. (1999). No effect of glucagon-like peptide-1 on short-term satiety and energy intake in man. The British journal of nutrition 81, 273–279.

46. Smits, M.M., and Holst, J.J. (2023). Endogenous glucagon-like peptide (GLP)-1 as alternative for GLP-1 receptor agonists: Could this work and how? Diabetes Metab Res Rev, e3699. 10.1002/dmrr.3699.

